# Comparison of WHO laboratory-based and non-laboratory-based CVD risk Charts among Hypertensive Adults Attending Primary Healthcare Centers in West Africa Sub-region

**DOI:** 10.1101/2025.01.05.25320020

**Authors:** Kojo Awotwi Hutton-Mensah, Olayinka Rasheed Ibrahim, Adaku M Nwankwo, George Bediako Nketia, Funmi Temidayo Adeniyi, Abukari Yakubu Natogmah, James Ayodele Ogunmodede, Dike Ojji, Olumide Adesola, Biodun Sulyman Alabi, Olugbenga Ayodeji Mokuolu, Daniel Sarpong

**Affiliations:** Directorate of Medicine, Komfo Anokye Teaching Hospital, Kumasi, Ghana; Department of Pediatrics, Division of Clinical Medicine, University of Global Health Equity, Kigali, Rwanda; Department of Internal Medicine, Gwarimpa General Hospital, Abuja, Nigeria; Department of Family Medicine/Polyclinic, Korle Bu Teaching Hospital, Ghana; Department of Pediatrics, University College Hospital, Ibadan, Nigeria; St Anthony Ann Hospital, Donyina; Department of Internal Medicine, University of Ilorin, Ilorin, Nigeria; Department of Internal Medicine, University of Abuja Teaching Hospital, FCT, Nigeria; Institute of Child Health, University of Ibadan, Ibadan, Nigeria; Department of ENT, University of Ilorin, Ilorin, Nigeria; Department of Pediatrics, University of Ilorin, Ilorin, Nigeria; Office of Health Equity Research, Yale School of Medicine

**Keywords:** Hypertension, WHO CVD risk charts, West-Africa subregion, Adults

## Abstract

**Background:** The World Health Organization (WHO) non-laboratory cardiovascular disease (CVD) risk chart is sub-region-specific and is advocated in resource-constrained settings. However, the extent of agreement with laboratory-based assessment among hypertensive adults attending primary health centers (PHCs) in the West Africa sub-region remains unknown. This study compared 10-year CVD risk among adults with hypertension attending PHCs in Ghana and Nigeria.

**Materials and Methods:** This cross-sectional study recruited 319 adults with hypertension at PHCs in Ghana and Nigeria. All participants had their blood pressures, anthropometrics, fasting blood sugar, and fasting cholesterol measured following standard procedures. WHO laboratory and non-laboratory CVD risks were assessed and compared using Kappa’s statistics, correlation, and Bland-Altman Plot.

**Results:** The median (interquartile range) for laboratory-based and non-laboratory-based CVD risk scores were comparable [7.0 (4.0 11.0) vs. 7.0 (4.0 to 11.0), p = 0.914]. Of the 319 participants, laboratory-based assessment classified 214 (67.1%) as low risk, while 210 (65.8%) were classified as low risk using the non-laboratory method. Eleven (3.4%) and 14 (4.4%) participants were classified as high-risk using laboratory- and non-laboratory-based methods, respectively. Overall, there was a very good positive correlation between the CVD risk assessment methods (r = 0.948, p<0.001). For all participants combined, there was substantial agreement (Kappa statistics), with K = 0.766. Bland-Altman showed a mean bias of 0.15 (SD = 1.74) in favor of non-laboratory-based assessment of CVD with an upper limit of 3.57 and a lower limit of -3.26.

**Conclusion:** There was substantial agreement between laboratory- and non-laboratory-based WHO CVD risk charts in this study. In low-resource settings, such as Ghana and Nigeria, the WHO non-laboratory CVD risk prediction model offers a huge opportunity for primary CVD prevention in adults with hypertension.

## Introduction

Cardiovascular disease (CVD) is the leading cause of death worldwide, accounting for 33% of all deaths with low- and middle-income countries like Ghana and Nigeria, accounting for as many as 80% of CVD-related deaths ^1^. In addition, the majority of CVD-related burden in Africa occurs in people between the ages of 30-70 years, and the resulting disability-adjusted life years (DALYs) poses serious economic and social consequences for families and the nation at large ^2, 3^. Consistent with global statistics, risk factors for CVD in Africa include hypertension, diabetes mellitus (DM), dyslipidemia, central obesity, tobacco use, physical inactivity, unhealthy diet, and hyperuricemia, most of which are preventable ^4^.

Considering the huge burden of CVD, as well as health inequity in Africa, emphasis must be placed on CVD disease prevention. Early detection is critical in disease prevention; hence, using CVD risk prediction tools will play a significant role in CVD prevention. There are many CVD risk models and scores that have been developed, including the WHO CVD risk score, Framingham risk score, pooled cohort equation and systematic coronary risk evaluation, among others ^5^. The sub-region-specific WHO CVD risk score chart has been advocated for use in West Africa to predict the 10-year risk of fatal and nonfatal CVD outcomes. It consists of non-laboratory (office-based) and laboratory-based charts ^6^. The laboratory-based chart utilizes information on age, sex, smoking status, systolic blood pressure (SBP), history or evidence of DM, and total cholesterol. In contrast, the non-laboratory-based chart includes information on age, sex, smoking status, SBP, and body mass index (BMI). Due to resource limitations in low and middle income countries, WHO non-laboratory CVD charts are favored, especially at primary healthcare levels where access to laboratory tests is limited ^7^. Although the data for the derivation of the WHO-CVD risk charts were from various sub-region data, including the West Africa sub-region, the assessment of the model performance has been advocated for various sub-populations ^8^. Thus, a comparison between laboratory and non-laboratory charts in the Eastern Africa sub-region showed that the latter underperformed in the diabetic sub-population ^9^. Similarly, in the Iran sub-population, in a high-risk group (score > 20%), there was limited agreement between the WHO-CVD laboratory and non-laboratory-based methods ^7^. At present, with an estimated population of 460 million and a hypertensive prevalence of 27%, the extent of agreement between WHO laboratory and non-laboratory risk among West Africans with hypertension remains unknown ^10,11^. Therefore, in this paper, we compared the WHO non-laboratory-based CVD risk score with the WHO laboratory-based risk score among patients with hypertension attending PHCs in Ghana and Nigeria.

## Materials and Methods

### Study design and settings

This cross-sectional descriptive study included adults with hypertension attending PHCs in Ghana and Nigeria. This study was conducted at the Okelele Primary Healthcare Center, Ilorin, Nigeria, and St. Anthony Ann Hospital, Donyina, Ashanti Region, Ghana. The two primary healthcare centers provided outpatient treatment and follow-up for adults with hypertension.

### Study participants

This study included patients aged 40–74 years with hypertension who consented to participate. Hypertension was defined as those already on treatment (controlled or uncontrolled), treatment-naïve with a blood pressure ≥ 140/90 mmHg, and newly diagnosed with three resting blood pressure measurements (BP≥ 140/90 mmHg). Adults with evidence of CVD (stroke, heart failure, peripheral artery disease, or ischemic heart disease) and pregnant women were excluded.

### Sample size estimation

Using a prevalence of hypertension of 27% in the West Africa sub-region ^10^, at a 5% level of precision and a 95% confidence interval, we estimated a minimum sample size of 300 (http://www.raosoft.com/samplesize.html).

### Recruitment and Data collection

A total of 319 participants were consecutively recruited between 1^st^ July 2023 and 28^th^ December 2023, with 160 participants from Nigeria and 159 from Ghana. We used a pretested questionnaire to gather relevant sociodemographic and cardiovascular risk factors from participants. Each participant underwent physical measurements, including blood pressure (BP) measurements.

### Physical measurements

Anthropometric measurements were performed according to the WHO guidelines. The weight was measured using an Omron HN286 electronic human weighing scale with an accuracy of 0.1 kg. Each participant’s height was measured with a “Seca 213” mobile stadiometer with an accuracy of 0.1 cm. BMI was calculated using the following formula: BMI (kg/m2) = weight (kg) / height (m^2^).

We measured each participant’s BP using a validated upper-arm BP monitor (‘Omron M7 Intelli IT’). In brief, each participant sat quietly with their feet on the floor and their clothes loosened around the arm for at least five minutes before blood pressure readings were taken. The displaced BP readings were documented in the study protocol. We obtained three serial BP measurements and three minutes apart, and used the average of the last two readings as a measure of BP values the data analysis.

### Cardiovascular disease risk score assessment

We calculated the CVD risk scores of participants using WHO CVD (laboratory-based) prediction charts and non-laboratory-based charts (2019 revised edition) ^6^. For laboratory-based prediction scores, each participant’s score was calculated based on age, sex, smoking status, presence or absence of diabetes, SBP, and total cholesterol. In the non-laboratory group, each participant had CVD risk scores based on age, sex, smoking status, SBP, and BMI.

### Outcomes

Comparison of laboratory-based CVD risk scores and non-laboratory-based risk scores in adults with hypertension attending PHCs in Ghana and Nigeria

### Data analysis

We analyzed the data from the study protocol using IBM SPSS version 29. Descriptive statistics were used to summarize participants’ sociodemographic variables. The WHO CVD risk scores did not follow a normal distribution and were presented as medians with interquartile ranges (IQR) and further compared using the Mann-Whitney U test. The continuous variables that were normally distributed (systolic BP, diastolic BP, cholesterol, and BMI) were compared using the independent T-test. Pearson’s correlation coefficients were used to assess the relationship between the two CVD risk score assessment methods. To assess the concordance and agreement between the two methods, we used Kappa statistics and Bland-Alman plots, respectively. The Kappa statistics were interpreted as follows: a < 0 indicated (less than chance agreement); 0.01 to 0.20 (slight agreement); 0.21 to 0.40 (fair agreement); 0.41 to 0.60 (moderate agreement); 0.61 to 0.80 (substantial agreement); and 0.81 to 0.99 (almost perfect agreement). The WHO CVD risk, which classifies the 10-year risk of CVD events, included risk scores < 5%, 5% to < 10%, 10% to < 20%, 20% to < 30%, and ≥ 30%, which indicated very low-, low-, moderate-, high-, and very high-risk groups, respectively. The level of statistical significance was set at p < 0.05.

### Ethical approval

This study was approved by the Ghana Health Service Ethics Review Committee (GHS-ERC:007/05/23) and the Kwara State Ethical Review Committee (ERC/MOH/2023/02/090). We also sought permission from the appropriate authorities at both primary healthcare facilities. A detailed explanation of what the study entails in information sheets, including study procedures, was made available to all participants in the language they best understood, and written informed consent was obtained. The data collected were coded to ensure the anonymity of the study participants and stored in a password-encrypted computer.

## Results

### General characteristics

A total of 319 adults with hypertension receiving care at PHCs in the two countries (160 from Nigeria and 159 from Ghana) participated in this study. The mean (standard deviation) age of participants was 59.1 (10.2) years. This study included 259 women (81.2%). Twenty-three (7.2%) participants had diabetes mellitus, and five smoked cigarettes (Table 1). The median (interquartile range-IQR) for laboratory-based CVD risk scores was 7.0 (4.0 to 11.0) and was comparable to the non-laboratory-based CVD risk scores (p = 0.914) (Table 1).

**Table 1:** General characteristics of the study participants.

| Variables | Total | Males<br>(n=60) | Females<br>(n=259) | P values |
| --- | --- | --- | --- | --- |
| Age group [Years] |  |  |  |  |
| ≤ 50 | 74 (23.2) | 15 | 59 | 0.931* |
| 51-60 | 94 (29.5) | 17 | 77 |  |
| >60 | 151 (47.3) | 28 | 123 |  |
| Smoke cigarette | 5 (1.6) | 3 | 2 | 0.048f** |
| Alcohol intake | 19 (6.0) | 12 | 7 | <0.001f** |
| Diabetes Mellitus | 23 (7.2) | 5 | 18 | 0.781f** |
| Systolic BP | 142.8 (24.4) | 144.4 (24.3) | 142.4 (24.4) | 0.579*** |
| Diastolic BP | 86.4 (13.1) | 86.2 (13.9) | 86.4 (14.0) | 0.914*** |
| Cholesterol | 5.2 (1.1) | 4.7 (1.0) | 5.3 (1.1) | <0.001*** |
| BMI [Kg/m<sup>2</sup>] | 26.5 (5.6) | 25.2 (4.9) | 26.8 (5.7) | 0.039*** |
| Laboratory-based | 7.0 (4.0 to 11.0) | 7.0 (4.0 to 13.8) | 7.0 (4.0 to 11.0) | 0.937# |
| CVD risk scores |  |  |  |  |
| Non-lab. based | 7.0 (4.0 to 11.0) | 7.0 (4.0 to 12.0) | 7.0 (3.0 to 11.0) | 0.923 |
| CVD risk scores |  |  |  |  |

BP-Blood pressure; BMI-Body mass index; CVD-cardiovascular disease, \*Chi-square;
\*\*Fischer exact test,\*\*\* T-test, # Mann-Whitney U test.

### CVD risk scores classification based on the laboratory and non-laboratory assessments

Of the 319 participants in this study, laboratory-based assessment classified 214 (67.1%) as low risk, while 210 (65.8%) were classified as low risk using the non-laboratory method. Eleven participants (3.4%) were classified as high-risk using laboratory-based assessment, whereas 14 (4.4%) were classified as high-risk using non-laboratory-based methods (Figure 1).

**Figure 1:**
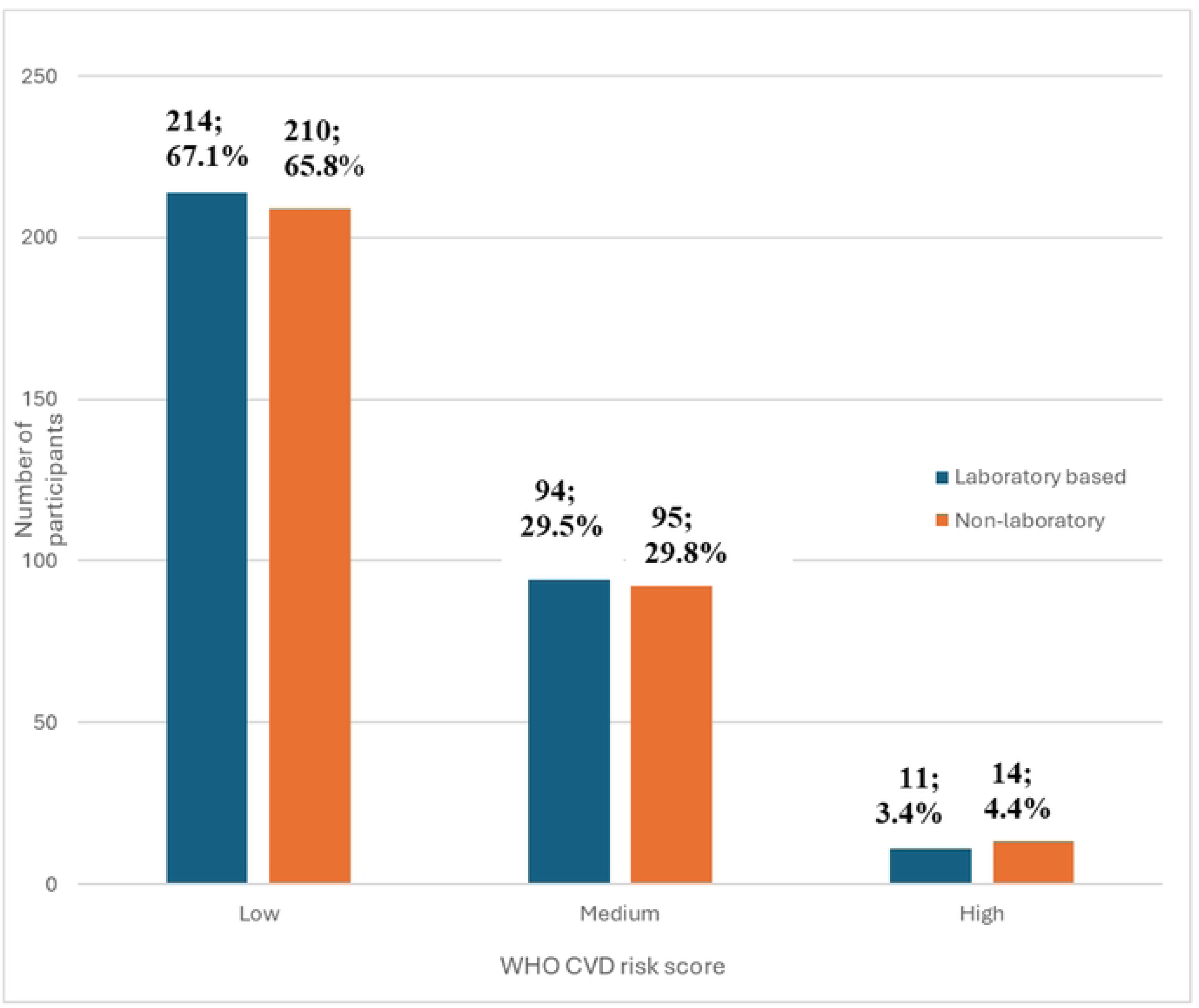
Classification of CVD risks an1ong the study participants.

### Correlation of the CVD risk scores between laboratory and non-laboratory assessments in the study participants

Overall, there was a strong positive correlation between the CVD risk assessment methods (r = 0.948, p<0.001). Based on the age stratification, participants aged 50 and below had a good positive correlation. In contrast, those older than 50 had a strong positive correlation between laboratory and non-laboratory-based assessments (Table 2).

**Table 2:** Correlation of the CVD risk scores between Laboratory and non-laboratory assessments in the study participants.

| Variables | Total | *r* | 95% CI | p |
| --- | --- | --- | --- | --- |
| **Age groups [Years]** |  |  |  |  |
| **40-74** | 319 | 0.948 | 0.936, 0.958 | <0.001 |
| **≤ 50** | 74 | 0.716 | 0.583, 0.812 | <0.001 |
| **51-60** | 94 | 0.868 | 0.807, 0.910 | <0.001 |
| **>60** | 151 | 0.893 | 0.855, 0.921 | <0.001 |
| **Sex** |  |  |  |  |
| **Male** | 60 | 0.938 | 0.898, 0.963 | <0.001 |
| **Female** | 259 | 0.952 | 0.939, 0.962 | <0.001 |

### Agreement between the laboratory and non-laboratory-based risk scores

For all participants combined, there was substantial agreement (Kappa statistics), with K = 0.766. Similarly, in both males and females, the kappa statistics showed good agreement (Table 3). Bland-Altman shows a mean bias of 0.15 (SD = 1.74) in favor of non-laboratory-based assessment of CVD with an upper limit of 3.57 and a lower limit of -3.26 [Figure 2].

**Figure 2:**
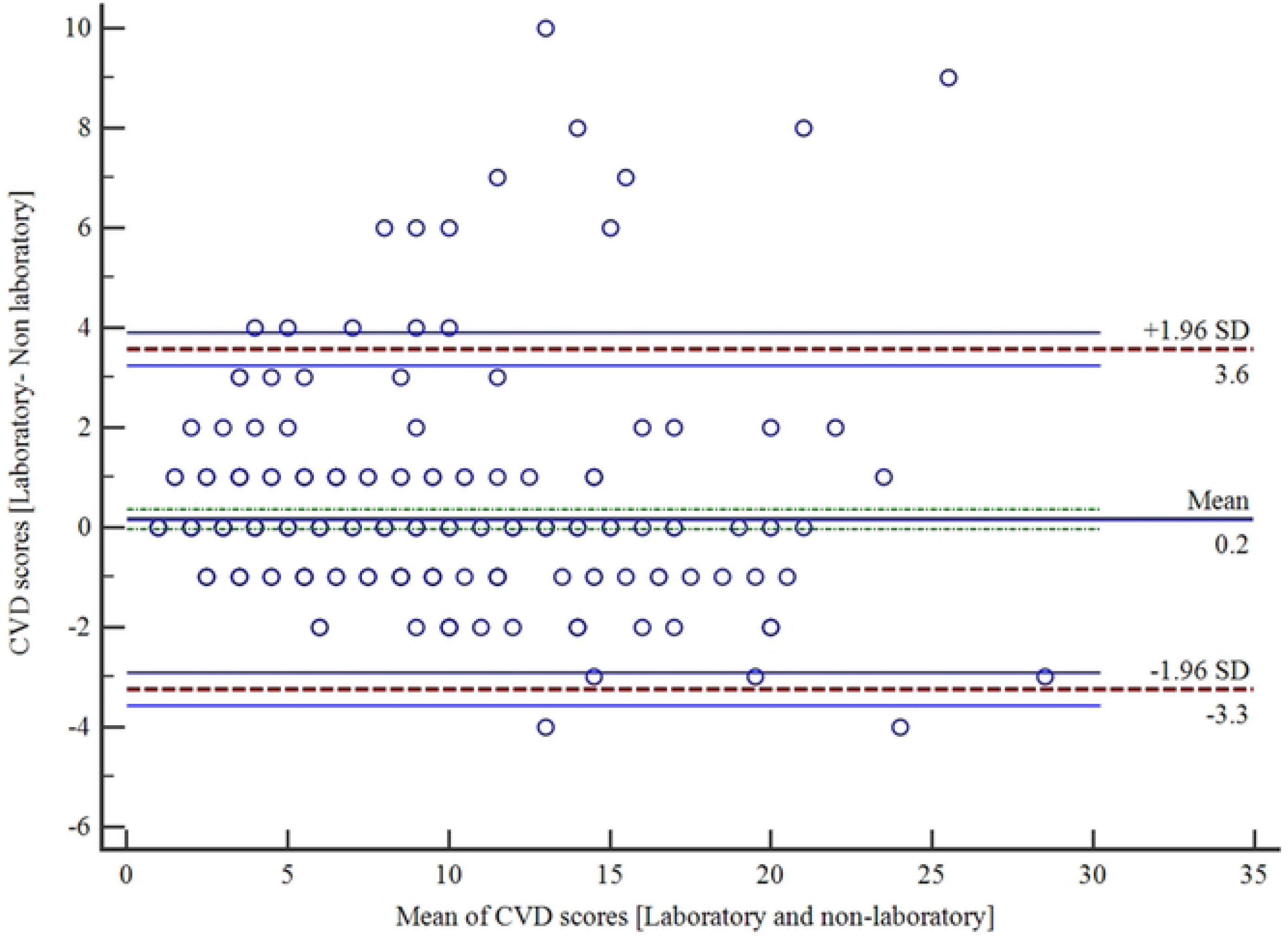
Bland-Altman plot showing agreement between the laboratory and non­laboratory assessment of CVD risk scores.

**Table 3:** Agreement between laboratory and non-laboratory-based risk scores.

| **Variables** | **Non-laboratory-based CVD risk** |  |  |  |  |
| --- | --- | --- | --- | --- | --- |
| **Laboratory-based CVD risk (All)** | **low** | **moderate** | **High** | **Total** | **Kappa (SE)** |
| **All** |  |  |  |  |  |
| Low | 198 | 16 | 0 | 21 | 0.766 |
| Moderate | 12 | 77 | 5 | 94 | (0.037), |
| High | 0 | 2 | 9 | 11 | p<0.001 |
| Total | 210 | 95 | 14 | 319 |  |
| **Male** |  |  |  |  |  |
| Low | 35 | 2 | 0 | 37 | 0.775 |
| Moderate | 3 | 14 | 1 | 18 | (0.079), |
| High | 0 | 1 | 4 | 5 | p<0.001 |
| Total | 38 | 17 | 5 | 60 |  |
| **Female** |  |  |  |  |  |
| Low | 163 | 14 | 0 | 177 | 0.763 |
| Moderate | 9 | 63 | 4 | 76 | (0.041), |
| High | 0 | 1 | 5 | 6 | p<0.001 |
| Total | 172 | 78 | 9 | 256 |  |

SE-Standard error.

## Discussion

The rising burden of CVD coupled with low health expenditure in sub-Saharan Africa remains a critical challenge that requires effective intervention ^1^. One key strategy to effectively combat CVD is to reduce its incidence through robust and effective screening aimed at identifying those at risk of adverse outcomes using CVD risk prediction models at primary healthcare levels, which are closer to the community. In this study, we compared the non-laboratory-based WHO CVD risk chart with the laboratory-based WHO CVD risk chart in adults attending PHCs in Nigeria and Ghana.

Most participants in this study were at ‘low risk’, with a median risk score of 7% for both laboratory- and non-laboratory-based. This finding is similar to other studies in Bhutan ^12^, India ^13^, Peru ^14^, and East Africa ^9^, where the average CVD risk scores ranged from very low-risk to low-risk among the studied cohort. However, in contrast to our findings, the average CVD risk score obtained in a study in Bangladesh was 10.3%, suggesting a moderate CVD risk ^15^. The differences between our study and that of Bangladesh may be due to variations in lifestyle, genetics, and socioeconomic factors in different populations ^10^. Our present study also shows that the non-laboratory risk chart predicted a slightly higher number of participants at high risk compared with laboratory-based assessment (4.4% vs. 3.4%), which is similar to a study in Bhutan, where the non-laboratory-based chart predicted 0.39% of participants as high risk compared to 0.23% as per the laboratory-based chart ^12^. This may be explained by the fact that both studies had a low prevalence of diabetes and smoking, which has been identified to impact risk stratification using non-laboratory methods significantly. This justification is further corroborated by a study in the East African sub-region, which showed that non-laboratory-based risk charts are suitable for people without diabetes in low-resource settings ^9^.

Our study showed substantial agreement between the two risk charts for both sex and age, with kappa statistics of 0.76, corresponding to substantial agreement. According to global work conducted by the WHO, there was a moderate agreement between the WHO 2019 CVD risk score using laboratory and non-laboratory algorithms ^6^. The findings of this study are comparable to findings from the ‘Fasa’ cohort study in Iran ^7^ as well as a cross-sectional study in Bhutan ^12^, where there was substantial agreement between the two risk scores with a kappa of 0.68 and 0.74, respectively, and among the North Indian population with a kappa statistic of 0.64 ^13^. The substantial agreement obtained in this study is similar to the substantial agreement of 0.78 obtained among people with normoglycemia and impaired fasting glucose in East Africa ^9^. However, the same study observed a moderate agreement of 0.46 in the Eastern African cohort with diabetes ^9^. The differences with respect to the diabetes group in East Africa may be due to the low number of adults with diabetes in this study (n = 23), and hence the minimal impact on the overall agreement between the two methods. The substantial agreement between the laboratory-based WHO CVD risk and the non-laboratory-based risk assessment has great implications for primary prevention of CVD in low-resource settings, such as the West Africa sub-region with low health expenditure per capita. Thus, this study supports using a simple and inexpensive non-laboratory-based WHO CVD risk chart for CVD risk screening at PHCs in the West Africa sub-region. This will facilitate the identification of those at high risk among hypertensive adults and ensure prompt early referral to higher levels for intervention.

In this study, the Bland-Altman analysis showed a mean bias of 0.15 in favor of the non-laboratory risk chart and the 95% limits of agreement ranging from -3.26 to 3.57, which denotes a reasonable agreement. The positive mean bias close to zero implies that the two methods generally agree, with minimal systematic differences. With a 95% confidence interval, including 0 implies minimal systematic difference (0.15), suggesting the difference is not statistically significant, and the methods agree. Our findings compare favorably with those of a study in Bangladesh ^15^ and an East African cohort study ^9^. While we did not perform a sub-analysis to explore the extent of agreement among sub-populations in this study, the limits of agreement obtained suggest that among hypertensive adults in Nigeria and Ghana, non-laboratory methods can be used in place of laboratory methods. Our study also showed a strong positive correlation (r = 0.948) between laboratory- and non-laboratory-based WHO CVD risk charts. The correlation was better in participants who were greater than 50 years old than in those who were less than 50 (0.880 vs. 0.716). This finding is corroborated by a population-based study using in Iran, where there was a robust correlation between laboratory-based and non-laboratory tests, with the correlation coefficient being higher in the elderly ^16^. A strong positive correlation between laboratory and non-laboratory WHO CVD risk charts underscore the accuracy of non-laboratory-based risk charts in predicting CVD risk. This provides an opportunity to address the unmet need for inexpensive and readily available risk prediction models for estimating the 10-year CVD risk in people seeking medical care at PHCs.

This is the first multi-country study in the West African sub-region to examine and compare laboratory-based and non-laboratory CVD risk charts at primary healthcare levels, where many of the populace receive care. Other strengths of this study include using multiple and appropriate statistical methods to compare the two risk charts, including the Kappa statistic, Bland–Attman plots, and correlation. Despite these strengths, this study had some limitations. The sample size might limit the generalization of the findings to Ghanaian and Nigerian populations. In addition, this study was cross-sectional; hence, causality could not be established, and participants were not followed up to see how many later developed CVD.

## Conclusion

The present study highlighted that CVD risk among patients with hypertension attending PHCs in Ghana and Nigeria is low, using the WHO CVD risk chart. This study observed substantial agreement between laboratory- and non-laboratory-based WHO CVD risk assessment. Therefore, in low-resource settings, such as Ghana and Nigeria, using a cost-effective and easily accessible non-laboratory CVD risk prediction model in PHCs offers a huge opportunity for primary CVD prevention in patients with hypertension. Using non-laboratory methods at PHC levels will ensure timely identification of patients at high risk of CVD, prompt referral for necessary interventions, and a subsequent reduction in the burden of CVD. Also, the non-laboratory method could also be used as a first line of screening for CVD risk among adults with hypertension and no diabetes mellitus. We encourage more extensive studies that include a significant number of people with DM to evaluate the generalizability of our findings to the general population.

## Data Availability

All relevant data are within the manuscript and its Supporting Information files.

## Acknowledgements

We thank the staff of Okelele primary health centre, Ilorin,Nigeria and St. Anthony Ann hospital, Donyina, Ghana for their contribution toward this study

## Notes

### Competing Interest Statement

The authors have declared no competing interest.

### Funding Statement

The author(s) received no specific funding for this work.

### Author Declarations

This study was approved by the Ghana Health Service Ethics Review Committee (GHS-ERC:007/05/23) and the Kwara State Ethical Review Committee (ERC/MOH/2023/02/090)

